# Predicting Residual Function in Hemodialysis and Hemodiafiltration – A Population Kinetic, Decision Analytic Approach

**DOI:** 10.1101/19001222

**Authors:** Mohammad I Achakzai, Christos Argyropoulos, Maria-Eleni Roumelioti

## Abstract

In this study, we introduce a novel framework for the estimation of residual renal function (RRF), based on the population compartmental kinetic behavior of Beta 2 Microglobulin (B2M) and its dialytic removal. Using this model, we simulated a large cohort of patients with various levels of RRF receiving either conventional high-flux hemodialysis or on-line hemodiafiltration. These simulations were used to estimate a novel population kinetic (PK) equation for RRF (PK-RRF) that was validated in an external public dataset of real patients. We assessed the performance of the resulting equation(s) against their ability to estimate urea clearance using cross-validation. Our equations derived entirely from computer simulations and advanced statistical modeling, and had extremely high discrimination (AUC 0.888 – 0.909) when applied to a human dataset of measurements of RRF. A clearance-based equation that utilized pre and post dialysis B2M measurements, patient weight, treatment duration and ultrafiltration had higher discrimination than an equation previously derived in humans. Furthermore, the derived equations appeared to have higher clinical usefulness as assessed by Decision Curve Analysis, potentially supporting decisions that for individualizing dialysis frequency in patients with preserved RRF.

## 1. Introduction

Patients with End Stage Renal Disease (ESRD) initiate dialysis when their intrinsic, residual renal function (RRF) is between 5-11 ml/min/1.73m2 [1,2]. Although RRF will be invariably lost within the first few years of dialysis initiation, sustaining this RRF is clinically important for the following reasons: low or declining RRF is associated with worse survival [3–7], worse phosphate control [8], increasing left ventricular hypertrophy [9], and poorer quality of life [3,10]. Hence existing dialysis guidelines [11,12] recommend the incorporation of RRF measurements into individualized patient prescriptions in order to achieve minimum dialysis adequacy targets.

In recent years, the paradigm of incremental dialysis [6,13,14], i.e. the gradual increase in frequency of dialysis match the RRF has also been proposed. This practice has not been widely adopted despite the fact that it may be associated with neutral[15] or even improved survival, reduced hospitalizations and improved control of several biochemical parameters[16] [17]. A major barrier to the safe implementation of incremental dialysis is the need to measure RRF in order to prevent underdialysis. This is a concern raised by regulators as well. In the United States the Center for Medicare Services explicitly considers twice weekly dialysis to be inadequate in patients with Urea Clearance (UrCl, a proxy for RRF) lower than 2 ml/min. Current guidelines [18] thus suggest that RRF be measured by interdialytic urine collection and plasma sampling for urea and creatinine for the calculation of the relevant clearances. However such collections are inconvenient for the patient [17,19–21], costly and partially adhered to, even in incentivized research settings [22].

To overcome the shortcomings of urine collections, effort has shifted towards estimating UrCl without having to collect urine. These efforts leverage pre dialysis measurements of “middle molecules”; such as beta 2 microglobulin, (B2M) [19,21,23], cystatin C (CysC) [21,23] and beta trace protein (BTP) [19,21], either alone or in combination, to estimate UrCl. These data suggest that B2M is the single most predictive biomarker of the guideline relevant cutoff of UrCl <2ml/min. Addition of other middle molecule markers e.g. CysC or BTP only marginally improves the performance of RRF estimating equations. Despite these encouraging results, these equations are not deployed in clinical practice because of the large variability in predicting the RRF of individual patients. This variable performance is thought to reflect the complex, multi-compartmental interdialytic kinetics of middle molecules and is particularly evident in patients with minimal or zero RRF [21].

In this study, we introduce a novel framework for the estimation of RRF, based on the population compartmental kinetic behavior of B2M and its removal during dialysis that have been meticulously modelled and meta-analyzed by our group [24,25]. This population kinetic model captures interindividual variability in the processes of generation, distribution and even elimination of B2M from the body. Using this model, we simulated a large cohort of patients with various levels of RRF receiving either hemodialysis (HD) or hemodiafiltration (HDF). These simulations were then used to estimate a novel population kinetic equation for RRF (PK-RRF) that was validated in an external public dataset of real patients [21]. Incorporation of additional renal function biomarkers in our approach is straightforward under the *simulator calibration* framework [26,27], that is commonly applied to calibrate simulations against real world measurements. This is an innovative direction towards the development of multi-biomarker models, which we explore in this manuscript. We assessed the performance of the resulting equation(s) against their ability to estimate UrCl using cross-validation. Clinical utility of these predictive models is quantified from a decision curve analysis/net-benefit perspective [28,29].

## 2. Experimental Section

The PK-RRF has two components: a) a statistical model linking B2M to RRF and b) a model that links RRF to a clinical measurement (urinary clearance) of a biomarker that could be used as a proxy for the RRF. The first component was developed by simulating the B2M kinetics in an artificial cohort of “patients” with characteristics that cover all possible combinations of dialysis prescriptions and kinetic parameters for B2M. After the conclusions of the simulations, a statistical description of the relationship between the output of the simulator (B2M) and the control parameters (e.g. dialytic clearance, B2M generation rate, distribution compartment volume, RRF) was derived. Such descriptions can be used to derive estimating equation for the actual RRF based on observable quantities that are either easily measurable (e.g. B2M), directly available (dialysis prescription), or can be proxied (e.g. dialytic clearance based on pre and post dialysis B2M levels).

After development, the estimating equation for the RRF was used as input to a second model that related the measured RRF to urinary clearance. The first component of the PK-RRF is derived by flexible parametric modeling of simulated data. The second component of the PK-RRF requires additional experimental data. In the context of this paper, key studies provided data that clarified the relationship between urinary clearance of urea and creatinine, and RRF. Even though the underlying population kinetic simulations and the emulator were based on sound principles, it would be naïve to expect them to faithfully represent the complexity of the B2M generation, distribution and removal by dialysis. Hence, we also consider calibrating the predictions of the PK-RRF against an external dataset of actual measurements performed in actual patients. Figure 1 presents a schematic overview of the approach adopted in this report.

**Figure 1.**
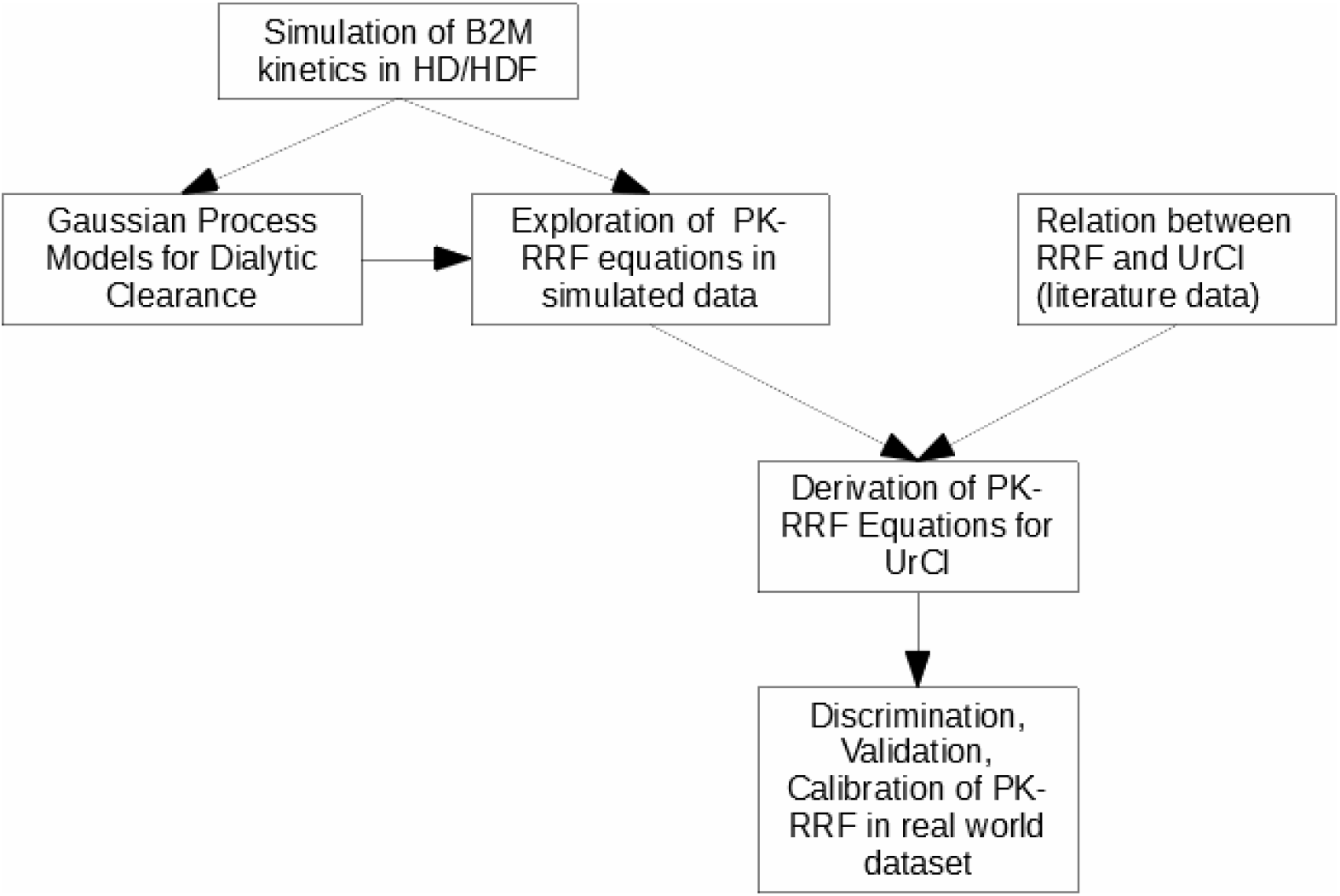
Overview of the approach adopted in this paper. We simulated the B2M kinetics in populations undergoing hemodialysis (HD) or hemodiafiltration (HDF). These simulations were used to derive Gaussian Process approximations for dialytic clearance and equations that related B2M to the known residual renal function (PK-RRF). Literature data were then used to relate the RRF to Urea Clearance (UrCl) and derive an equation that uses B2M and dialysis parameters to predict UrCl, a proxy for the unmeasured RRF. These in-silico derived equations were assessed in terms of discrimination, validation and calibration in real world datasets.

### 2.1. Simulations of B2M kinetics during dialysis and hemodiafiltration

We simulated 10,000 hemodialysis patients receiving either thrice weekly high flux HD or on line hemodiafiltration (HDF) under the two compartment, variable volume model for B2M kinetics [24,30] following the methodology previously reported by our group [24]. These simulations used the PopK parameters for B2M and the range of the dialyzer B2M clearances [24,31] described in our previous meta-analysis. Dialysis-related parameters (dialysis duration, ultrafiltration volume, substitution flow rates) were based on the FHN [32,33], HEMO [34] trials for the HD simulations and on the three largest randomized controlled trials (Dutch CONTRAST [35], Spanish ESHOL [36], Turkish OL-HDF [37]), for HDF. Each patient was given a unique RRF value and the interdialytic/intradialytic changes in B2M concentration were simulated over a period of three months with the LSODA integrator [38]. The pre dialysis and post dialysis simulated concentration of B2M at the first weekly session at the end of the three month period were extracted from the simulation files and were used for model development. This simulated dataset is available on-line as Table S1.

### 2.1. Development of PK-RRF equations

#### 2.1.1. Generalized Additive Models for the PK-RRF

The B2M pre dialysis values in the simulated dataset (Table S1) were used to predict the RRF value in each artificial patient, via means of Generalized Additive Models (GAM) [39,40]. GAM regressions allow the data-driven discovery of complex, possibly non-linear interactions between continuous covariates and either continuous (the RRF value) or discrete (RRF > 2 ml/min) outcomes. GAMs achieve these goals by modeling the relationships between variables through smoothing functions, e.g. thin plate regression splines (TPRS) or Gaussian Processes (GP). Contrary to other popular flexible models used in biomedicine e.g. cubic splines, TPRS can model highly complex relationships and nonlinear interactions between covariates and without the need to specify knot locations, i.e. ranges of values in which the relationship between the covariate and outcome changes mathematical form. For models with more than two interacting variables measured in different scales, we also considered GPs when fitting the GAMs. Despite their flexibility, GP models require a considerable amount of data to learn these relationships, and thus could only fit on the dataset of simulated patients. In fitting these models, one has to specify a covariance function that describes the correlation of the value of quantity modelled as GP at two different points of the input (e.g. B2M, dialyzer clearance, body weight, treatment duration) variables. For the purpose of this work, we used the Kamman and Wand version of the Matérn covariance function [41] in the mgcv GAM package in R.

#### 2.1.2. In silico exploration of factors affecting the performance of the PK-RRF

While exploring our artificial dataset, we developed a series of models that used control parameters of the simulations; e.g. dialytic clearance of B2M or treatment time in addition to B2M levels when predicting RRF. The purpose of this modeling was to understand the additional measurements or dialysis treatment parameters that should be incorporated into a basic model of B2M vs. RRF in order to develop an estimating equation that can truly individualize predictions. For these analyses, we adopted a hold-out validation strategy in which the simulated dataset was randomly split in a training/development (2/3 of all data) and a testing/validation subset. The simplified formulas were then developed in the training dataset and their performance was assessed in the development one. Dialytic clearance was calculated from treatment duration, ultrafiltration volume, body weight, pre and post dialysis B2M measurements via the Leypoldt formula [43]. This formula was obtained through a uni-compartmental approximation to the full kinetic model of B2M. The inputs to this formula are the pre and post dialysis B2M levels, body weight and ultrafiltration rate. It is not entirely clear how accurate this equation is over the entire range of the input parameters. Hence, we used our kinetic simulations to compare this formula against a GP that utilized the same variables using a hold out validation technique. These analyses were carried out in Microsoft R Open v3.4.0-3.5.1; GAM analyses were undertaken via the package mgcv.

### 2.2. Modeling the relation of RRF to UrCl

A key question that has not been answered in the literature [14,42,43], concerns the quantitative relationship between clinical measurements of CrCl and UrCl and their relationship with the concept of the RRF. In the context of this work, the RRF maps to the glomerular filtration rate (GFR), as has been recently discussed [42,42,44,45]. Only a few landmark, experimental studies [46–49] have simultaneously measured GFR using exogenous markers and timed urine collections, but the data from these studies have not been analyzed together. We thus undertook a quantitative synthesis of the relationship between GFR measurements and the timed urine collections for the calculation of clearances of urea and creatinine. In the compartmental kinetic model for B2M, the RRF participates in its non-normalized to the Body Surface Area form, ie. its units are expressed in ml/min rather than ml/min/1.73m2 and this is the form we analyzed in this paper. We extracted individual patient data (Table S2) from these publications from the relevant tables, or figures using digitizer software as previously described [25]. These studies had used a variety of methods; e.g iothalamate, inulin or DTPA, to measure GFR (mGFR). The UrCl and CrCl data from these studies were related to the RRF using the following hierarchical measurement error model:

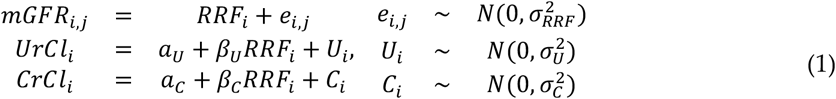

According to this model, the jth measurement of GFR (*mGFR*_*i,j*_ j=1,2,3 corresponding to iothalamate, inulin or DTPA) in the ith patient i an unbiased, but noisy estimate of the (unobserved) RRF in that patient (*RRF*_*i*_). The (observed) *UrCl*_*i*_ and *CrCl*_*i*_ in the same patient are linearly related to the *RRF*_*i*_, with the relationship incorporating linear intercept (*α*_*U*_ and*α*_*C*_, respectively) and slope (*β*_*U*_ and*β*_*C*_, respectively terms. If the urine clearances are unbiased estimates of the RRF, the corresponding intercept terms should be zero; if the slope parameters are less (greater) than 1.0, the clearance measurements consistently under(over)estimate the RRF. The quantities (*e*_*i,j*_, *U*_*i*_ and *C*_*i*_) are random terms, quantifying measurement and formula error in the measured GFR, UrCl and CrCl respectively. All random terms are modelled as zero mean, Gaussian (normal) random variables with standard deviations given by the sigma symbols in (1). Model fitting procedures are discussed in Appendix A.

### 2.3. Measurements of B2M, RRF and dialyzer clearance in patients

We utilized the publicly available dataset of RRF, CysC, B2M, urea, creatinine and dialysis parameters that accompanied the publication of Vilar et al [21] to evaluate the PK-RRF equation performance. This dataset included measurements in 341 individuals, of whom 230 were receiving HDF and the remaining conventional, high flux HD. Thirty six per cent of the participants in this dataset had RRF of 0 ml/min, providing an opportunity to assess equation performance in this challenging subgroup of patients. For the purpose of this paper we used the 24 hour urea clearance as an index of RRF. This is the approach taken by the bulk of the literature to date [17,19,21,23] in the field. Post dialysis B2M, and thus dialytic clearance calculations were available in 291 patients in the Vilar cohort, thus allowing us to fit the dialytic clearance equations.

### 2.4. Models and outcomes for UrCl

We compared our PK-RRF equations against the Shafi equation for the a) primary outcome of having a UrCl > 2 ml/min vs ≤ 2 ml/min and b) the continuous prediction of UrCl. In the publication describing the Shafi equation, the formula’s continuous prediction was thresholded in order to classify patients as having a clearance above or below the cutoff. This is also how we used the formula in this paper. The Shafi equation was developed in a cohort of 44 patients with urine volume >250ml undergoing HD and were validated in a cohort of 826 mixed cohort of patients undergoing either peritoneal dialysis or HD from the NECOSAD study. In the PK-RRF approach we developed logistic GAM regression model for the threshold of RRF corresponding to a UrCl ≤ 2 ml/min. We considered two logistic PK-RRF equations as comparators of the Shafi equation: i) a *basic equation* that included only the predialysis B2M, and ii) a *clearance based* equation which used a GP to account for dialytic clearance as explained in section 2.1.2..

### 2.5 Model Comparison: Discrimination, Calibration and Clinical Usefulness

We adopted the AUC for discrimination of UrCl ≥ 2ml/min vs UrCl< 2ml/min (the cutoff of incremental dialysis in the existing guidelines) as the primary metric for evaluating the performance of the logistic PK-RRFs against the Shafi equation. Other descriptive metrics were also computed to provide a more complete evaluation for the continuous UrCl outcome: the proportion of predictions within a 0.5 – 2 ml/min of the true RRF, measures of bias (Mean Absolute Error, Median Error), variance (Interquartile Range) and the total Root Mean Square Error (RMSE) between the model predictions and the actual RRF measurements.

The basic method for assessing calibration was the *linear calibration plot*. In these analyses, the UrCl, or the linear predictor for the logistic PK-RRF was used as a covariate in linear regression analyses against the real world measurements [50,51]. This linear regression yields an intercept and a slope which can be used used to assess calibration. The intercept is an index of the formulas ability to be systematically too high or low (“calibration-in-the-large”) and should be close to zero for an unbiased model. The calibration slope should be ideally equal to unity, with values smaller than one reflect model overfit, with the departure from unity quantifying the effects of overfit.

#### 2.5.1 Clinical Usefulness (Decision Curve Analysis)

We also assessed the clinical usefulness of the PK-RRF and Shafi models using Decision Curve Analysis (DCA). DCA essentially quantifies the net numbers of correct classifications gained by applying a predictive model or a simple clinical decision rule and the resulting clinical consequences of a treatment decision guided by the rule. DCA [28] assumes that the threshold probability of a clinical state at which one would opt in or out of treatment decided by the prediction rule, is informative of the risk benefit ratio of a false-positive and a false-negative prediction. This relationship is then used to calculate the Net Benefit (NB) over different threshold probabilities contrasting the benefit against treatment strategies of treating all or no patients. The NB is a quantity that assumes values between negative infinity up to the prevalence of the predicted state with positive values indicating that the model had a beneficial impact on clinical decision-making and negative values indicating harm. The Standardized Net Benefit (SNB) indexes the NB to the prevalence of the predicted state, to give a maximum potential value of one. Since there is not usually a single, universally acceptable probability threshold, one can plot SNB against threshold probability to obtain a “decision curve”. DCA then identifies the magnitude of benefit, and allows a direct comparison of several models against the range of threshold probabilities, or equivalently risk-benefit ratios. For any given risk threshold, the DCA shows the prediction model with the highest utility [52]. In our context, we applied DCA under the opt-out treatment policy framework [53]: the PK-RRF and the clinical rule based on the Shafi formula are used to identify low risk patients (patients with preserved UrCl > 2 ml/min) who could opt out of the (reference) treatment strategy of thrice weekly dialysis.

### 2.6 PK-RRF recalibration and inclusion of multiple biomarkers

Finally we explored the potential of re-calibration to improve the PK-RRF performance; in particular, we carried out analyses of “internal-external” leave-one-out cross-validation [54] of the PK-RRF against the Vilar dataset. In these analyses, we sequentially set aside each of the original study participants and repeat the linear calibration regression analysis. After these models have been fit, we generate predictions for each patient held back and compare the model prediction against the actual RRF measurement. We also examined the possibility of linear calibration of the PK-RRF equations using multiple additional biomarkers (urea, creatinine and CysC). To do so, we applied the “simulation-calibration” framework proposed from the statistical literature about computer simulations [26,27,55]. This framework was put forward to calibrate simplified versions (our PK-RRF equations) of complex computer models (such as the output of our kinetic simulations) against real world measurements of the phenomena (B2M levels – RRF). The statistical framework for the calibration analysis rests on *Gaussian Process* models, thus providing a unique methodological synthesis between the GAM models used to develop the PK-RRF and this calibration analysis. For the purpose of this work, we assessed whether prediction models that included statistical interactions among the measurements of multiple biomarkers (B2M, CysC, urea and creatinine) and the PK-RRF can be used to improve discrimination and calibration of the PK-RRF equation against the Vilar dataset. Whereas these models do include the same slope and intercept terms as the simple linear calibration models, they differ by incorporating GPs among the biomarkers that could predict RRF. These additional terms, effect a non-linear correction of the B2M based equations as they allow the contribution of the B2M measurement to non-linearly co-vary along with measurements of other biomarkers of renal function when predicting UrCl.

### 2.6. Software Availability

Due to the complexity of the TPRS models, it is not easy to write down the equation in a mathematically simple form, like other equations. We thus distribute the entire R code required to estimate the PK-RRF from the software repository https://bitbucket.org/chrisarg/pk-rrf/.This software repository includes all code required to fit the models considered in this paper. It also includes code to set up a *shiny* application to deploy these equations in a web server. An instance of this server application has also been deployed at https://chrisarg.shinyapps.io/popk_rrf/. This server version is only intended for demonstration purposes (e.g. due to the licensing of the *shiny* platform, it will only be running for 25 cumulative hours every month).

## 3. Results

### 3.1. Development of PK-RRF and performance of GP for dialytic clearance

Use of a pre dialysis level B2M achieved very high discrimination for predicting ≥ 2 ml/min (AUC 0.896). Inclusion of postdialysis time and dialytic clearance improved discrimination (AUC 0.915) in the simulated dataset (Appendix A, Figure A1). When benchmarked against the true value, the dialytic clearance calculated on the basis of the Leypoldt formula had a median (mean) bias of - 0.78 (−3.82) ml/min and highly variable performance: an IQR (standard deviation) of the difference between true and estimated values of 14.6 (22.50) ml/min. On the other hand the dialytic clearance estimated by the GP had minimal median (mean) bias of 0.18 (0.07) ml/min and much less variable performance: the IQR (standard deviations) of the difference between true and estimated value of 8.29 (7.21) ml/min (Appendix A, Figure A2). In summary, the analysis of the simulated dataset suggests that one should include not just B2M, but also GP based estimates of dialytic clearance in order to derive a predictive equation for RRF that has high discrimination.

### 3.2. The relationship between RRF, Urea and Creatinine Clearance

The literature data suggest that UrCl and CrCl are linearly related to the mGFR (Figure 2), while regression estimates are shown in Table A1.

**Figure 2.**
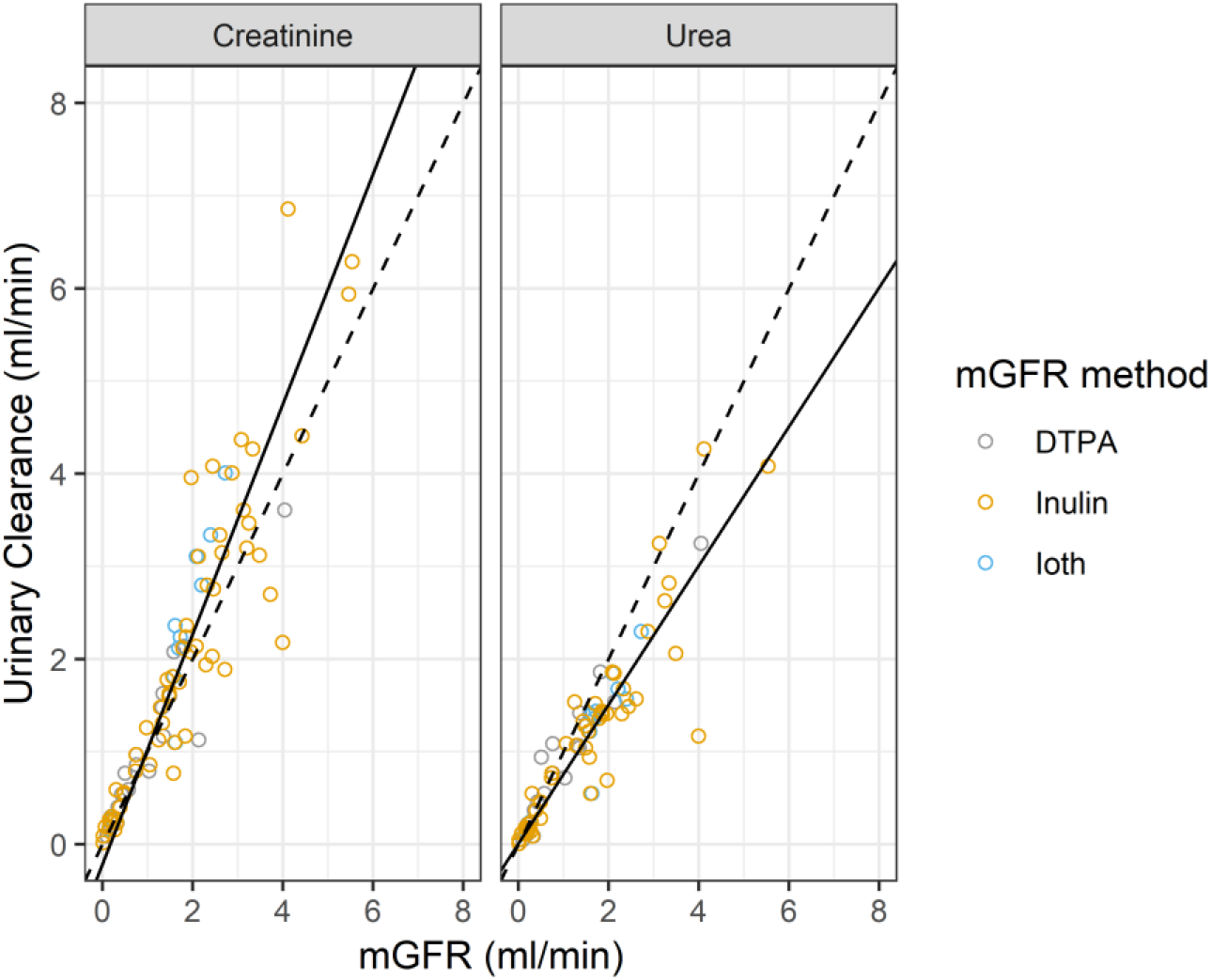
Relationship between measured GFR (mGFR)/RRF and urinary clearance for Urea and Creatinine in the literature data. Dashed line: line of identity with zero intercept (bias) and unity slope. Continuous lines are the results of the regression analysis based on Eq (1). See also Table A1.

Urea clearance measurements had essentially zero bias in predicting mGFR (the intercept of the linear regression was 0.007 ml/min), while underestimating mGFR (the regression slope was 0.751). There was a small negative bias when CrCl is used to estimate mGFR, yet CrCl systematically overestimated mGFR (its slope was 1.242). These analyses suggest the following simplified equation to relate UrCl to RRF:

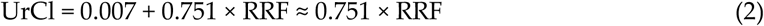

In the latter equation ignoring the intercept and the uncertainty in the parameter estimates is justified because of the small value of the former parameter and the extremely high signal to noise ratio in estimating the slope. In particular, the 95% credible interval for the slope is between 0.694-0.810, indicating that we are 95% certain that the true value of this parameter is contained in this narrow range. In all subsequent analyses, we multiplied the output of the PK-RRF estimating equations for the continuous RRF outcome by 0.751 to convert it to an estimate for the UrCl. Furthermore, when deriving PK-RRF equations that predict the odds of having UrCl above a given cutoff, e.g. 2 ml/min, one has to use the equivalent RRF cutoff of 2/0.751 ml/min ∼ 2.66 ml/min.

### 3.3. The logistic PK-RRF has high discrimination and calibration when validated against a patient dataset

The PK-RRF equation developed in simulations, achieved greater discrimination (AUC 0.829 – 0.911) compared to the Shafi equation in the entire Vilar cohort, the subset of patients on HD or HDF (Table 1). Discrimination by either equation was lower when applied in the entire dataset, i.e. not excluding anuric patients.

**Table 1.** AUCs for predicting RRF > 2 ml/min in the entire Vilar cohort

| Subset | Equation | HD | HDF | All |
| --- | --- | --- | --- | --- |
| Non Anuric | PK-RRF | 0.845 | 0.823 | 0.829 |
|  | Shafi | 0.807 | 0.782 | 0.787 |
| All Patients | PK-RRF | 0.911 | 0.878 | 0.888 |
|  | Shafi | 0.842 | 0.816 | 0.821 |

HD: Hemodialysis, HDF: Hemodiafiltration

Inclusion of treatment duration, body weight, predialysis and postdialysis B2M as GP increased the performance of the PK-RRF equation (AUC 0.909 for all patients and 0.855 for the non-anuric ones). The p-values of the Spiegelhalter test for overall calibration accuracy of prediction probabilities were 0.594 (basic model), and 0.743 (*clearance based* model). Examination of the calibration plots (Appendix C, Figure C1) showed that all these models systematically underestimated the risk of having UrCl ≤2 ml/min (intercept different from zero). Nevertheless, the clearance based model had a calibration slope of 1.058, close to the ideal value of unity, indicating that the model’s variables and their interconnection validly generalize from the simulated to the real world dataset. Other metrics (e.g. Brier score, Somer’s rank correlation, unreliability index) favored the clearance based PK-RRF over the basic one (Figure C1). These analyses suggest that an “intercept update”*[56,57]* of the PK-RRF to improve “calibration-at-large” may allow the use of these models to human populations with different overall probabilities for the outcome of UrCl ≤2 ml/min. The superior calibration of the *clearance based* PK-RRF was also shown when the RRF was examined as continuous outcome (Appendix C).

### 3.4. Re-calibration of the PK-RRF and incorporation of multiple biomarkers

We explored the possibility of improving the performance of the PK-RRF equations using recalibration and additional biomarkers. Towards that goal we crossvalidated the logistic and continuous RRF equations in all patients with CysC, pre dialysis urea and creatinine measurements in the Vilar dataset. Recalibration using multiple biomarkers improved the discrimination of the clearance-based RRF compared to a simple intercept/slope recalibration that did not use these markers (Table 2). Overall, the performance of the PK-RRF equation improved but only marginally (delta AUC was ∼ 0.05 – 0.1) with the inclusion of multiple biomarkers. Across the entire dataset of patients receiving either HD or HDF, incorporation of the pre dialysis urea and creatinine had similar incremental improvement as the inclusion of CysC.

**Table 2.** Discrimination of recalibrated clearance-based PK-RRF for predicting UrCl > 2 ml/min

| Additional Biomarkers | HD | HDF | All |
| --- | --- | --- | --- |
| None | 0.868 | 0.922 | 0.900 |
| Creatinine | 0.888 | 0.922 | 0.904 |
| Urea | 0.885 | 0.929 | 0.906 |
| Cystatin C | 0.866 | 0.927 | 0.903 |
| Cystatin C + Urea | 0.881 | 0.927 | 0.906 |
| Creatinine + Urea | 0.887 | 0.927 | 0.907 |
| Cystatin C + Creatinine | 0.878 | 0.924 | 0.903 |
| Cystatin C + Creatinine + Urea | 0.856 | 0.938 | 0.907 |

HD: Hemodialysis, HDF: Hemodiafiltration.

Examination of the proportion of the predicted UrCl within a fixed amount (in ml/min) for the continuous UrCl (Figure 3) reproduced the patterns seen for the discrete outcome. Analysis of bias, variance and total error (Table 3) for the Cystatin C and the Urea/Creatinine calibrated models are shown in Table 3. Overall, incorporation of additional biomarkers appears to improve performance but only marginally.

**Table 3.** Bias, Variance, Total Error and Precision of the continuous RRF outcome

| Metric | Equation | HD | HDF | All |
| --- | --- | --- | --- | --- |
| Median<br>(ml/min) | Clearance based PK-RRF | 0.09 | 0.23 | 0.18 |
|  | Clearance based PK-RRF+Urea/Creatinine | -0.14 | 0.23 | 0.14 |
|  | Clearance based PK-RRF+Cystatin C | -0.13 | 0.22 | 0.10 |
| MAE<br>(ml/min) | Clearance based PK-RRF | 0.93 | 0.88 | 0.89 |
|  | Clearance based PK-RRF+Urea/Creatinine | 0.86 | 0.83 | 0.84 |
|  | Clearance based PK-RRF+Cystatin C | 0.91 | 0.82 | 0.85 |
| IQR<br>(ml/min) | Clearance based PK-RRF | 1.37 | 1.03 | 1.19 |
|  | Clearance based PK-RRF+Urea/Creatinine | 1.11 | 1.02 | 1.12 |
|  | Clearance based PK-RRF+Cystatin C | 1.25 | 1.06 | 1.15 |
| RMSE | Clearance based PK-RRF | 1.26 | 1.25 | 1.25 |
| (ml/min) | Clearance based PK-RRF+Urea/Creatinine | 1.14 | 1.16 | 1.15 |
|  | Clearance based PK-RRF+Cystatin C | 1.20 | 1.17 | 1.18 |

HD: Hemodialysis, HDF: Hemodiafiltration, MAE : Mean Absolute Error, IQR: Interquartile Range, RMSE: Root Mean Square Error

**Figure 3.**
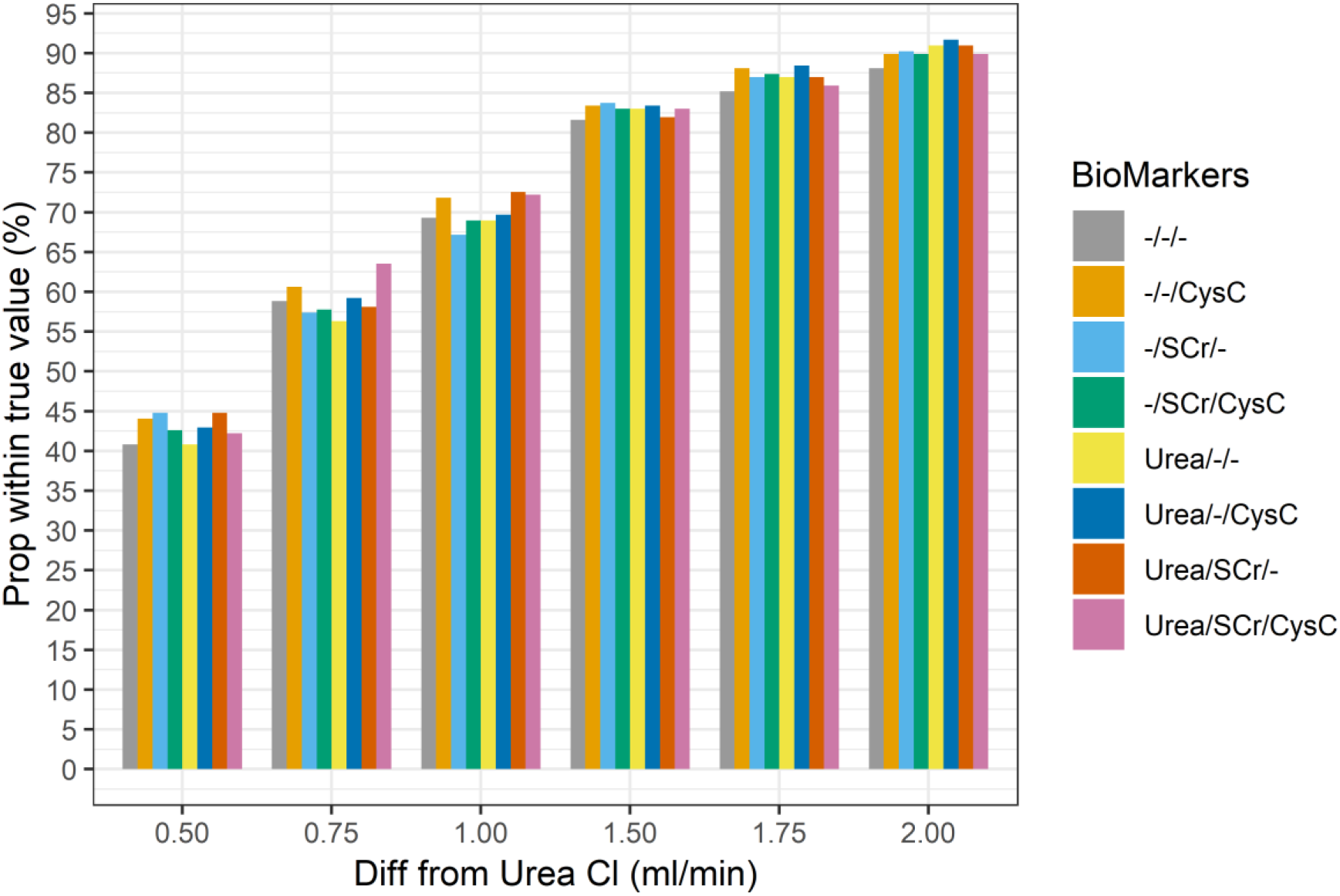
Proportion of predicted UrCl within a certain fixed amount from the measured UrCl for calibrated models that included additional biomarkers

### 3.5. Clinical Usefulness (Decision Curve Analysis)

We investigated the clinical usefulness of the PK-RRF equations in detecting patients with UrCl > 2 ml/min (“low risk group”) who could be offered infrequent or incremental dialysis in the Vilar dataset. Decision curves were constructed for the entire range of threshold probabilities from 0 to 1, corresponding to increasing risk benefit ratio from 1:100 to 100:1. SNB were calculated for the base PK-RRF, the clearance based PK-RRF and the Shafi clinical decision rule (Figure 4, left). The default strategy of offering incremental/infrequent dialysis has by default a SNB of zero irrespective of the probability threshold used to classify patients as low risk (gray horizontal line). The Shafi rule was dominated by both the base and the clearance base PK-RRF when the risks of offering patients the possibility to opt out of the default strategy were much smaller than the perceived benefit. In particular, the SNB of both PK-RRF rules were less than −0.1 when the threshold for classifying patients as low risk were less than 0.28. The Shafi rule dominated the base PK-RRF between threshold values of 0.28 – 0.85 and dominated by it for higher thresholds, corresponding to a higher perceived risk than the anticipated benefit. The clearance base PK-RRF had similar performance as the Shafi equation in the mid-range of threshold probabilities and dominated it at either low or high values. The PK-RRF models and the Shafi rule dominated a strategy of treating no patients with the default policy.

**Figure 4.**
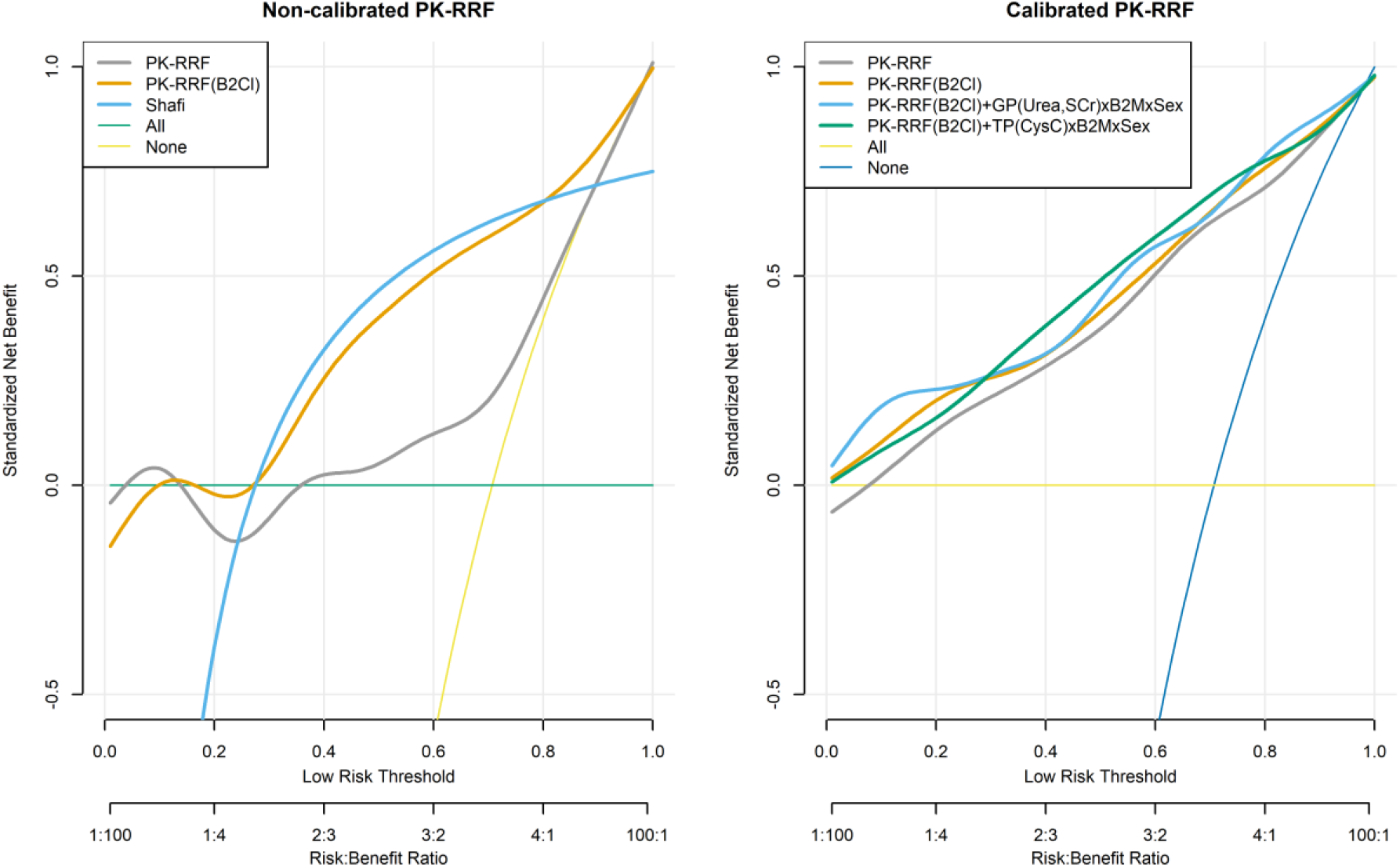
Clinical Usefulness (Decision Curve Analysis) for predicting the binary outcome of UrCl>2ml/min and to offer twice weekly or incremental dialysis. Left Graph: uncalibrated base PK-RRF model (“PK-RRF”), clearance based PK-RRF model (“PK-RRF(B2Cl)”), Shafi clinical decision rule (“Shafi”), treat “all” with thrice weekly dialysis, treat “none” with thrice weekly. Right graph: recalibrated basic and clearance based PK-RRF, clearance-time based PK-RRF incorporating urea and creatinine (interacting with the B2M measurement and gender) and the clearance based PK-RRF model that incorporates the Cystatin C. All PK-RRF curves were smoothed using a non-parametric smoother prior to plotting.

Finally, we assessed the potential improvement in clinical usefulness of the recalibrated base and clearance PK-RRF and the inclusion of additional biomarkers. These analyses were performed under a leave-one out cross validation framework and are shown in the right panel of Figure 4. As expected, linear re-calibration improved the performance of the base PK-RRF model much more than the clearance-based model alone. Inclusion of urea and creatinine did not appear to improve the clinical usefulness of the clearance-based PK-RRF model, except for low risk thresholds. On the other hand, inclusion of CysC measurements resulted in models with higher clinical usefulness in the middle range of threshold probabilities.

## 4. Discussion

In this paper we utilized a novel population compartmental kinetic framework to derive a set of equations for the prediction of RRF in patients undergoing conventional high flux HD or on line HDF. Our equations were derived entirely with computer simulations and advanced statistical modeling, and had extremely high discrimination when applied to a human dataset of measurements of RRF. A clearance-based equation that utilized pre and post dialysis B2M measurements, patient weight, treatment duration and ultrafiltration had higher discrimination than a equation previously derived in humans. Furthermore, the derived equations appear to have higher clinical usefulness supporting decisions that depend on patients having preserved RRF e.g. incremental dialysis.

Compartmental models for biomarker kinetics are familiar to nephrologists since they have been used to quantify dialysis dose for decades [58–63]. These models are mathematical descriptions of the processes of generation, distribution, and elimination that determine the concentration of the biomarker of interest. A population viewpoint extends the kinetic approach by allowing interindividual variation in these parameters. This interindividual variation allowed us to simulate the relation between B2M, RRF and the impact of the dialytic regimen in a manner that generalized from the simulated patients to the real world clinical data. The resulting equations, which were entirely derived in artificial datasets, thus, had high discrimination when applied to data from actual patients. In fact the performance of the PK-RRF equations rivalled the performance of equations derived entirely in human populations e.g. AUC of 0.91 [21] and 0.84 [23]. Overall our paper adds to the expanding literature showing that plasma levels of middle molecules in general and B2M in particular [19,21,23] can predict the regulatory relevant threshold of RRF > 2ml/min on a par with the performance of the clinically accepted, validated troponin assays in the diagnosis of acute coronary syndromes (AUC: 0.84-0.94) [64].

The high performance of the PK-RRF equations are entirely due to the validity of the constructs used to derive them from first principles. These constructs include the bi-compartmental kinetics of the B2M, the population distribution [24] of the kinetic parameters of B2M, the effects of dialytic clearance [25] and finally the relation between the RRF and the clinical measurement (UrCl) used as its proxy. Despite the high theoretical validity of our approach, translation of the derived equations to the real world should be expected to not be entirely free of complications. In fact, we have documented the need for calibration of the base and the clearance-based PK-RRF formulas. The degree of calibration required to predict RRF in a new dataset appears to be smaller than that required to adapt the high quality Shafi equation to the same external dataset. Recalibration of the PK-RRF equations did not materially affect their extremely high discrimination but did seem to have a positive impact on the clinical usefulness as assessed by DCA. Consequently, we feel that the calibrated version of the base PK-RRF equation should be used over the uncalibrated version. However, the uncalibrated clearance-based PK-RRF equation appears to perform equally well and either the calibrated or the non-calibrated version can be applied to future clinical studies.

Despite the success of middle molecules in predicting RRF, a puzzling feature of the literature to date [19,21,23] concerned the marginal success of multi-biomarker equations in the field. This was also noted in our study, which showed unimpressive improvements in discrimination and precision when CysC or simultaneous urea and creatinine measurements were used to recalibrate the B2M based PK-RRF. Although there have been concerns that the involvement of B2M in the inflammatory response may confound the relationship between RRF and B2M [42,65], our approach to consider variable rates of generation in our large scale simulations probably allowed us to derive PK-RRF equations insensitive to large variations in the generation rate. Hence, our formulas are unlikely to require additional biomarker measurements for robust performance. Despite these observations, we have noticed that some improvement in clinical utility may be derived by considering additional, readily measurements e.g. urea, creatinine or CysC. We also provided the analytical methodology to incorporate these biomarkers, as flexible GP embedded in the recalibration framework. This opens the possibility of incorporating additional, promising biomarkers e.g. BTP [42,43] in datasets that have measured them.

In developing our equations we were motivated by the unmet need for measurements that can facilitate further research in the area of RRF preservation and/or implementation of incremental, individualized forms of dialysis in practice [13,43]. Research in both areas is impeded by the lack of alternative techniques to measure RRF that do not require urine collections [19,21,23,42]. Prospective observational [66], and retrospective propensity score matched studies [15] have shown that an incremental approach to dialysis frequency is non inferior with respect to mortality and may be associated with improved quality of life. Considering the direct treatment cost differential, i.e. biweekly dialysis has 2/3 dialytic costs than thrice weekly dialysis, a formula that can identify patients with relative preserved RRF could have direct implications for both research and practice of “personalized dialysis”. However, this research should take place within the boundaries of existing regulations for the dialysis industry. In fact, one of the barriers in practice incremental dialysis in the United States is a concern raised by regulators: the Center for Medicare Services explicitly considers twice-weekly dialysis to be inadequate in patients with RRF lower than 2 ml/min. By providing a PK-RRF equation that can predict this threshold with high discrimination and positive Net Benefit across the entire spectrum of the risk-benefit assessments, we feel that research in this space can proceed in an ethical and regulatory compliant manner.

A few limitations of the developed PK-RRF equations should be kept in mind. First, the analytical complexity precludes the ability to write them down in closed form, similar to the simpler equations they outperform. This is an unavoidable price to pay for the high discrimination of the PK-RRF equation. Nevertheless, we provide these equations as software programs in the open source R programming language and a web server in order to allow other investigators to replicate our results. Second, the formulas were validated only in cross-sectional assessments and their use in repeated evaluations of the RRF of the same patient remain untested. This is an area of exploration in future studies. Third, the marginal improvement of the discrimination of the multi-biomarker models may reflect deficiencies in the biomarkers available for inclusion. In particular, urea, creatinine and CysC, the conventional serum biomarkers of estimating renal function in patients not on dialysis via eGFR formulas, exhibit large interdialytic variation in levels and thus may not provide the optimal additional biomarkers. Fourth, the recalibrated PK-RRF equations have an intermediate validation status since we did not have an additional dataset to test their performance. This limitation does not extend to the uncalibrated version whose discrimination and clinical usefulness can be externally validated.

In summary, we have used computer simulations, the population kinetic approach and advanced statistical modeling to develop equations that can predict RRF (as assessed by UrCl) in patients undergoing maintenance HD or on line HDF. These equations exhibit high discrimination and clinical usefulness when validated against an external, public clinical dataset. Recalibrated versions of these equations were developed in a cross-validation setting and are available for clinical use as well. Future studies should validate these equations in repeated assessments of the same patients and explore the utility of the PK-RRF equations as research tools in the areas of preservation of RRF and incremental, personalized dialysis.

## Data Availability

All data and software code used in this paper are available from bitbucket

https://bitbucket.org/chrisarg/pk-rrf/src/master/

## Supplementary Materials

None

## Author Contributions

conceptualization, C.A. and M.E.R.; methodology, C.A.; software, C.A.; validation,

M.E.R. and M.I.A; formal analysis, C.A.; investigation, M.I.A and M.E.R.; data curation, M.E.R.; writing—original draft preparation, C.A.; writing—review and editing, M.I.A and M.E.R.; visualization, C.A.; supervision, C.A.; project administration, C.A.

## Funding

This study received no external funding.

## Acknowledgments

None

## Conflicts of Interest

“The authors declare no conflict of interest.”

## Appendix A: Development of PK-RRF equations and assessment of dialyzer clearance

For these analyses, we split the simulation dataset into a development dataset (n=13,333) and a validation (n=6,667) cohort. The basic PK-RRF emulator that incorporated only the B2M concentration achieved a moderate R2, and a high AUC (0.896) for classifying “patients” as having a RF> 2ml/min (Figure A1, panel “B2M Only”). There was substantial variation in predicting the RRF of individual “patients” as evidenced by an IQR of 4.1 ml/min, while precision was also small, since only 12.9% and 49% of predictions were within 0.5 and 2 ml/min respectively of the predicted values. Incorporation of the postdialysis dialysis level and its interaction with the predialysis level (panel “B2M+Post+B2M*Post”) did not materially improve precision or discrimination. Adding treatment time as a covariate (panel “B2M+Post+B2M*Post+T”) increased AUC to > 0.90. Nevertheless, inclusion of an index of dialyzer clearance and treatment time did improve the classification performance and increased the AUC to 0.915 (panel “B2M+Post+B2M*Post+T+Kd+T*Kd”).

**Figure A1.**
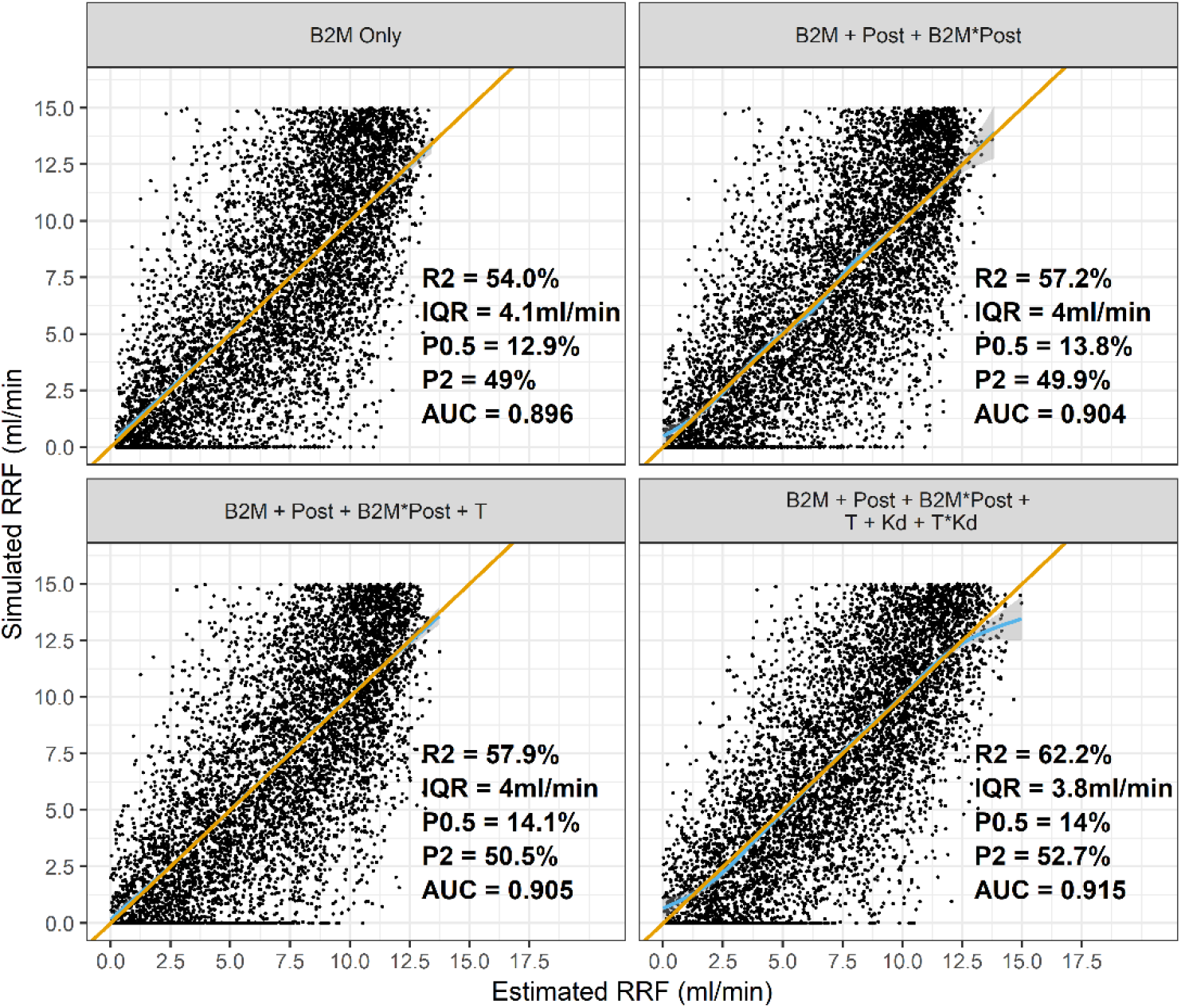
Indices of precision (P05/P02: probability of a prediction within 0.5 and 2 ml/min of the simulated RRF, IQR: interquartile range), model explanatory power (R2) and classification accuracy (AUC: Area Under the ROC curve) for predicting a RRF > 2ml/min across a range of models that included only the pre dialysis B2M concentration, the postdialysis level (Post), Dialysis Treatment Time (T), Dialysis Clearance (Kd). In models that incorporated statistical interactions between any two parameters, the interacting covariates are joined by the multiplication (“*”) symbols. Blue curve: smoothed average prediction, orange line: the line of identity, i.e. the ideal situation in which the prediction (y-axis) is identical to the simulated value (x-axis). These analyses were carried out in simulated datasets of 10,000 patients receiving hemodialysis and 10,000 receiving on-line hemodiafiltration.

In these analyses the value of the dialytic clearance was assumed to be known, a condition that does not apply in real world practice in which measurement of clearance does not take place. Hence we assessed the performance of the Leypoldt equation against a flexible, GP equation that used the same variables (body weight, ultrafiltration, pre and post dialysis B2M). These analyses are shown in Figure A2. While both approaches were associated with small (median) bias in the overall population, the performance of the GP was much less variable and separated the clearances between HD and HDF.

**Figure A2.**
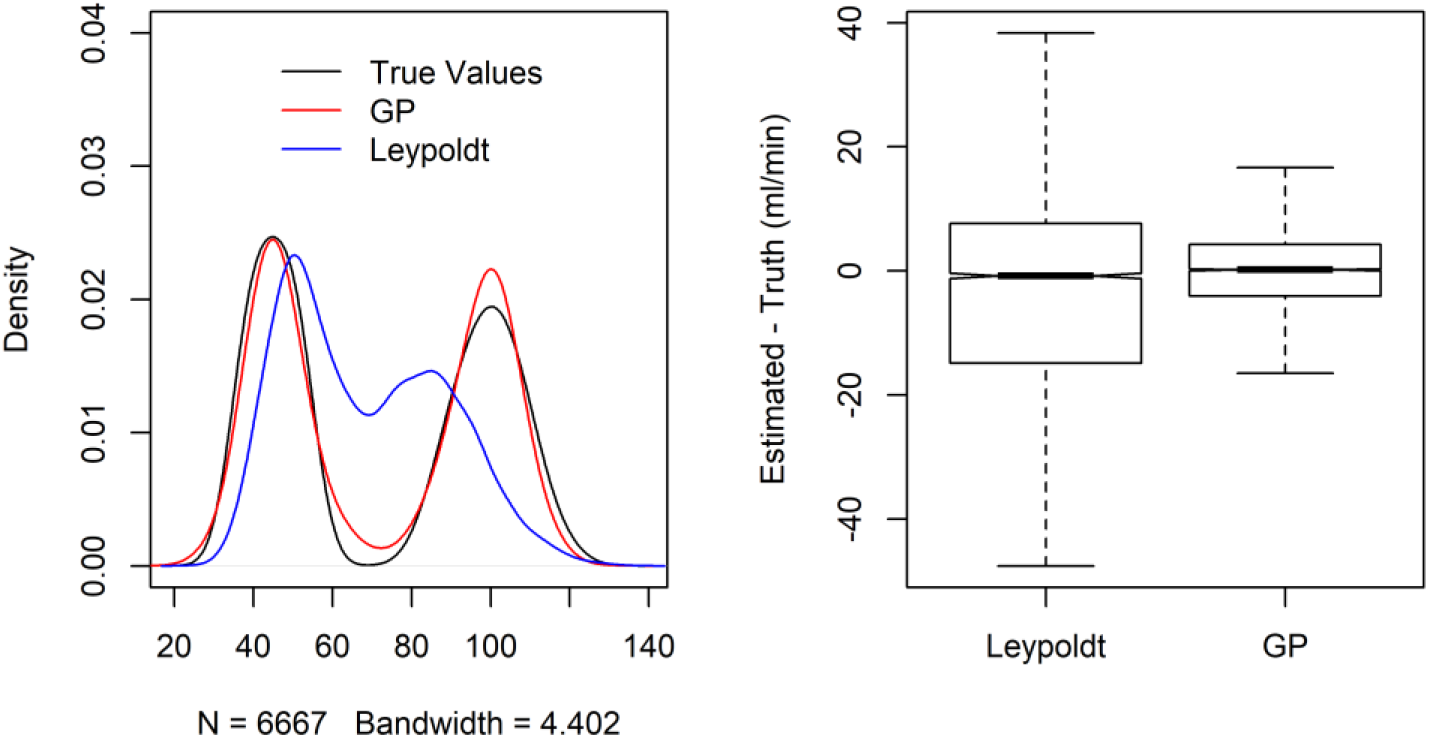
Smooth density histograms (left) of simulated and estimated dialytic clearances by the Leypoldt and GP approaches. The true distribution of clearance values is a bimodal one, because of the different clearances in hemodialysis and hemodiafiltration. The boxplots (right) show the differences between the estimated and simulated (true) values for the two methods. Overall the GP estimated clearance was much closer to the true one, as evidenced by the near complete superposition of the histogram of estimated values (red line, left graph) against the true values (black line, left graph), and the much tighter distribution of the differences between estimated and true values (boxplot, right graph)

## Appendix B: Fitting literature RRF and Urea/Creatinine Clearance Data

We fit the model of (1) to the literature data from a Bayesian perspective while assuming typical non-informative priors for all the unobserved quantities in the model. For the fitting we utilized long Markov Chain Monte Carlo (MCMC) simulations which were carried out with the *JAGS* software[67,68]. Technical parameters of MCMC runs were as follows: the Mersenne-Twister[69] was used as the underlying random number generator, number of independent chains (n=5), number of samples in the adaptive phase (n=100,000) discarded (burnin) samples (n=100,000), post-burnin samples (n=100,000), thinning interval (n=50) leading to 10,000 independent samples for each quantity of interest. Convergence of the MCMC was assessed by visual (trace) plots and the Gelman-Rubin diagnostic for multiple chains [70,71]. Although definitely an overkill, this long MCMC simulation ensured that the Potential Scale Reduction factors of the Gelman-Rubin diagnostic were indistinguishable from one (indicating successful convergence), and Monte Carlo (numerical) error was at most 0.001 for all parameters of interest. Means and standard deviations were used to summarize samples from the posterior simulations for the slope and intercept parameters, since their non-parametric kernel density estimates were visually perceived to be symmetric, gaussian-like densities. R/JAGS code and data are available in the online repository set up for this project. Model estimates are shown in Table A1. Monte Carlo numerical error was less than 0.001 for all parameters, while the potential scale reduction factors were one indicating successful convergence of the MCMC algorithm

**Table B1.** Estimates for the parameters of Equation (1) relating RRF to UrCl and CrCl

| Parameter | Mean | SE | Q2.5% | Q25% | Q50% | Q75% | Q97.5% |
| --- | --- | --- | --- | --- | --- | --- | --- |
| $\alpha_C$ | -0.213 | 0.124 | -0.458 | -0.296 | -0.213 | -0.130 | 0.033 |
| $\alpha_U$ | 0.007 | 0.072 | -0.135 | -0.040 | 0.007 | 0.056 | 0.150 |
| $\beta_C$ | 1.242 | 0.047 | 1.149 | 1.211 | 1.242 | 1.274 | 1.334 |
| $\beta_U$ | 0.751 | 0.030 | 0.694 | 0.731 | 0.751 | 0.770 | 0.810 |
| $\sigma_C$ | 0.564 | 0.080 | 0.414 | 0.510 | 0.561 | 0.615 | 0.728 |
| $\sigma_U$ | 0.316 | 0.048 | 0.229 | 0.284 | 0.314 | 0.346 | 0.416 |
| $\sigma_{RRF}$ | 0.302 | 0.046 | 0.223 | 0.269 | 0.299 | 0.331 | 0.401 |

## Appendix C: External validation of the PK-RRF in the Vilar dataset via linear calibration plots

A calibration analysis of the Shafi and clearance based PK-RRF equation against the measured RRF is summarized in Table C1. Calibration analysis was conducted on the enitre Vilar cohort of anuric and nonanuric patients. The PK-RRF demonstrated a small and statisticallyno significant amount bias in both HD and HDF. The Shafi equation had considerable bias (0.79 ml/min) in the HD subgroup. Calibration slopes for both models were different from the ideal value of one, yet they were closer to unity for the PK-RRF model. These analyses indicate the need for model re-calibration for both equations.

**Table C1.** Linear Calibration analysis for models of the continuous RRF in the Vilar cohort

| Parameter | Equation | HD | HDF | All |
| --- | --- | --- | --- | --- |
| Intercept | PK-RRF | 0.23 (0.19) | -0.12 (0.14) | 0.01 (0.11) |
| (ml/min) | Shafi | 0.79 (0.13) | 0.10 (0.11) | 0.44 (0.08) |
| Slope | PK-RRF | 0.57 (0.06) | 0.69 (0.05) | 0.64 (0.04) |
| (per 1 ml/min) | Shaffi | 0.36 (0.03) | 0.63 (0.04) | 0.48 (0.03) |

HD: Hemodialysis, HDF: Hemodiafiltration. Results are reported as model estimate (standard error).

**Figure C1:**
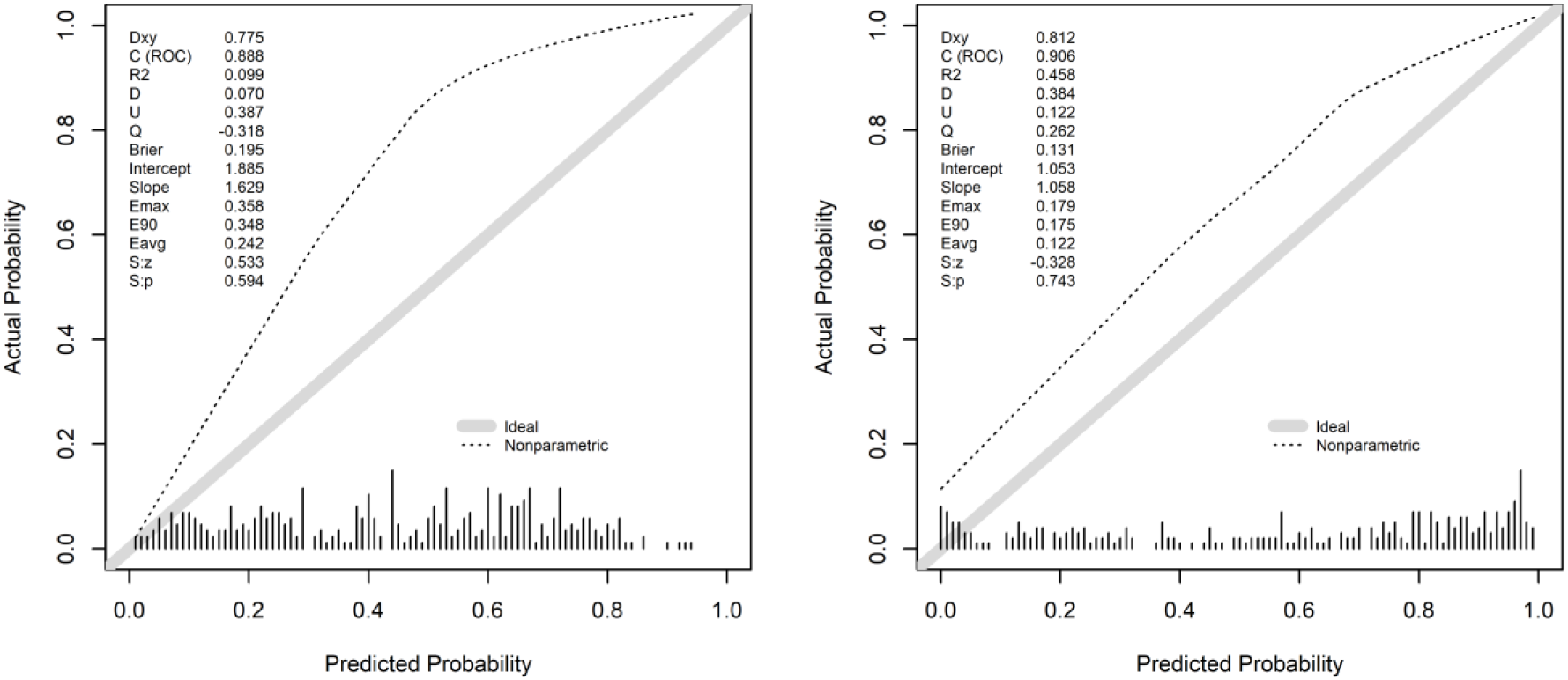
Calibration plots for the basic (left) and clearance based (right) logistic models. Dashed curve: non-parametric estimate of the calibration relationship between actual and predicted probability, grey line: ideal relationship (intercept of zero and slope of one). Dxy: Somer’s rank correlation, C(ROC): AUC for discrimination, R2: Nagelkerke-Cox-Snell-Maddala-Magee R-squared index, D: discrimination index, U: unreliability index, Q: quality index, Brier: Brier score (average squared difference in predicted and actual probabilities), Emax/E90/Eavg: Maximum/90^th^ quantile, average absolute difference in predicted and smoothed calibrated probabilities, S:z/S:p the z and two sided p-value of the Spiegelhalter test for calibration accuracy. Graphs generated by the “val.prob” function in the RMS R-package. Due to minor differences in algorithms, there is a difference in the third significant decimal from the AUC values reported in Table 1 of the main text.

## Notes

### Competing Interest Statement

The authors have declared no competing interest.

### Author Declarations

All relevant ethical guidelines have been followed and any necessary IRB and/or ethics committee approvals have been obtained.

Any clinical trials involved have been registered with an ICMJE-approved registry such as ClinicalTrials.gov and the trial ID is included in the manuscript.

